# Beyond BMI: Waist-Integrated BMI Improves Identification of Central Dysmetabolic Obesity

**DOI:** 10.1101/2025.07.07.25331012

**Authors:** Suresh N. Shinde, Rituparna S. Shinde, Succhietra Y. Bhangaaley

**Author notes:** **Corresponding author:** Dr. Suresh N. Shinde, MD., ‘Muktashree’, 5 Shelar Road, Bibwewadi, Pune, India 411037.

## Abstract

**Background:** Body mass index (BMI) does not capture fat distribution, particularly central adiposity, which is more closely linked to insulin resistance and cardiometabolic risk. We evaluated a waist-integrated anthropometric framework to identify dysmetabolic obesity beyond BMI.

**Methods:** In a retrospective observational study of 331 adults attending a metabolic clinic, an integrated body mass index (iBMI; weight × waist circumference/height) was derived. In a non-diabetic reference cohort, the relationship between iBMI and BMI was modeled, and deviation from this relationship was quantified. Associations among deviation metrics, insulin resistance, and visceral adiposity, as measured by bioelectrical impedance and dual-energy X-ray absorptiometry, were examined.

**Results:** Deviation from expected iBMI was significantly associated with visceral adiposity and insulin resistance, beyond BMI, and performed comparably to fasting glucose but better than glycated hemoglobin.

**Conclusions:** A waist-integrated BMI framework identifies central dysmetabolic obesity not captured by BMI and may enable earlier recognition of metabolic risk in clinical practice.

## Introduction

Body mass index (BMI) is widely used to classify overweight and obesity, but does not reflect fat distribution, particularly central adiposity, which is more closely linked to insulin resistance and cardiometabolic risk. Consequently, individuals with similar BMIs may have markedly different metabolic profiles, leading to risk misclassification across populations and body types. Waist circumference is a widely used surrogate for visceral adiposity, but when used alone, it lacks normalization for body size. To address these limitations, we propose an integrated body mass index (iBMI), defined as weight multiplied by waist circumference and normalized by height. By incorporating central adiposity and using height normalization without squared-height scaling, iBMI provides a more physiologically congruent representation of body geometry. Because absolute iBMI values vary by stature and population, interpretation requires a reference framework. We therefore derive an empirically defined relationship between iBMI and BMI in a non-diabetic cohort. Deviation from this relationship is hypothesized to reflect excess central adiposity, independent of body size.

## Methods

### Study design and population

This was a physician-led, retrospective, observational study conducted at a metabolic clinic in India. Clinical data from 331 adults attending routine outpatient consultations over a one-year period were analyzed. All data were anonymized before analysis. Ethical approval with an informed consent waiver was granted by the institutional ethics committee.

### Anthropometry and biochemistry

Body weight, height, and waist circumference were measured using standard clinical protocols. BMI was calculated as (weight/height^2^). An integrated body mass index (iBMI) was defined as weight × waist circumference/height. Fasting plasma glucose, fasting plasma insulin, and glycated hemoglobin (HbA1c) were measured using standard biochemical and immunoassay methods.

Insulin resistance and β-cell function were estimated using HOMA1-IR and HOMA1-B, respectively. (1) Visceral adiposity was estimated using bioelectrical impedance analysis. (2) In a subset of participants, visceral adipose tissue was also quantified using dual-energy X-ray absorptiometry. (3) A non-diabetic reference cohort was defined as individuals with fasting plasma glucose < 126 mg/dL and HbA1c < 6.5% (48 mmol/mol). Within this cohort, a linear regression analysis of iBMI and BMI was performed to define the adipose tissue equipoise line (EPL). Deviation from the EPL was quantified mathematically (observed iBMI minus ‘expected iBMI from the EPL’) as the iBMI residual (dBMI-res) and geometrically as the perpendicular distance to the EPL (graphical risk distance, GRD). Unsupervised k-means clustering was applied to fasting glucose, insulin, and visceral fat to identify metabolic phenotypes. Receiver operating characteristic analysis assessed discrimination for insulin resistance using a HOMA1-IR threshold of >1.7 (4). Analyses were conducted using Python.

## Results

Unsupervised clustering of fasting plasma glucose, fasting plasma insulin, and bioelectrical impedance–derived visceral fat identified three distinct metabolic phenotypes that spanned conventional BMI categories (Fig.1A). Principal component analysis showed progressive separation among clusters, reflecting increasing metabolic dysregulation.

**Figure 1.**
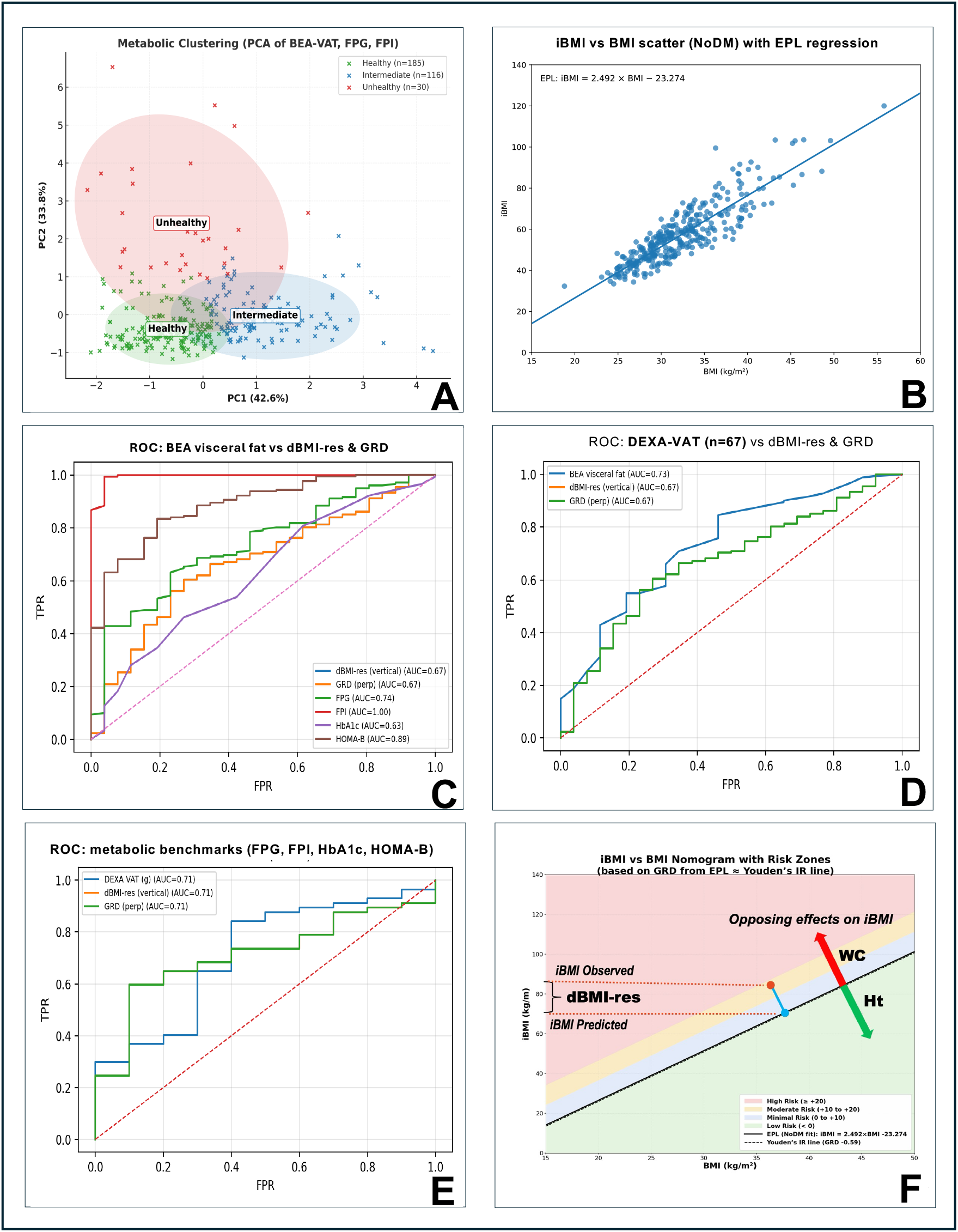
iBMI–BMI framework and metabolic risk stratification. (A) Principal component analysis identifying latent metabolic phenotypes. n = 331 (B) iBMI versus BMI relationship in a non-diabetic cohort with the adipose tissue Equipoise Line (EPL). n = 208 (C–E) Receiver operating characteristic analyses comparing vertical residual (dBMI-res) and perpendicular deviation (GRD) against metabolic and adiposity benchmarks. (F) iBMI–BMI nomogram illustrating GRD-based risk zoning relative to the EPL. iBMI, integrated body mass index; EPL, equipoise line; GRD, graphical risk distance; dBMI-res, observed iBMI − iBMI predicted from the EPL.

In a nondiabetic subset (n = 208), iBMI showed a strong linear relationship with BMI, yielding a regression line designated the adipose tissue equipoise line (EPL). The EPL represents the expected iBMI for a given BMI under conditions of preserved metabolic coupling between adiposity and insulin sensitivity. This regression served as the basis for deriving deviation metrics. The regression equation for the EPL, so derived, was iBMI = 2.492 × BMI − 23.274 (Fig.1B). Deviation from the EPL was quantified as the analytic residual (dBMI-res) and as the perpendicular geometric distance, designated the graphical risk distance (GRD).

In receiver operating characteristic analyses using HOMA1-IR > 1.7 to define insulin resistance, dBMI-res and GRD showed identical discrimination (AUC ≈ 0.67), with overlapping ROC curves across the full range of false-positive rates (Fig.1C). Their performance was comparable to that of fasting plasma glucose and exceeded that of glycated hemoglobin in this early-risk cohort.

Both dBMI-res and GRD were significantly associated with elevated visceral adiposity estimated by bioelectrical impedance (AUC ≈ 0.67) (Fig.1D). In a DEXA-imaged subset (n ≈ 67), similar associations were observed with imaging-derived visceral adipose tissue (AUC ≈ 0.67), confirming concordance with a reference modality (Fig.1E).

An iBMI–BMI nomogram incorporating the EPL illustrated graded risk zones, enabling visualization of deviation-based metabolic risk (Fig.1F). Receiver operating characteristic analysis identified an optimal GRD cut point using Youden’s index, reflecting the statistically optimal balance of sensitivity and specificity for insulin resistance and visceral fat burden. The cut point was close to zero, supporting GRD = 0 (corresponding to the EPL) as a physiologically interpretable reference.

## Discussion

This study introduces and validates a waist-integrated anthropometric framework that extends conventional BMI by explicitly accounting for central adiposity and anchoring interpretation to a physiological reference. By defining an empirically derived EPL in a non-diabetic population, we demonstrate that deviation from this reference captures clinically relevant dysmetabolic risk not adequately reflected by BMI alone.

Unsupervised metabolic clustering revealed distinct phenotypes that spanned conventional BMI categories, underscoring the dissociation between overall body mass and metabolic risk. Individuals classified as Metabolically Intermediate or Unhealthy by glucose, insulin, and visceral fat measures often overlapped with “normal” or modestly elevated BMI ranges. (5-7) The EPL represents the expected coupling between total body mass and central adiposity under metabolically compensated conditions. Individuals lying near this line appear to maintain a proportional distribution of adipose tissue, whereas deviations above the EPL reflect disproportionate central fat accumulation relative to body size. The analytic residual, dBMI-res, and GRD quantify this deviation as continuous signals. Their discriminatory performance, as confirmed by ROC analyses, is indistinguishable, indicating that both metrics capture the same underlying biological phenomenon.

The EPL was derived from a non-diabetic reference cohort. This design choice prioritizes physiological interpretability over causal inference, as longitudinal studies indicate that adiposity and glycemia become decoupled after the onset of overt diabetes due to catabolic weight loss and β-cell failure. (8,9)

In ROC analyses, deviation-based metrics showed moderate discrimination for insulin resistance, comparable to fasting glucose and superior to glycated hemoglobin in this early-risk cohort, yet remained inferior to insulin-based indices. This hierarchy is physiologically consistent: dBMI-res and GRD index upstream adipose distribution, whereas insulin and HOMA-derived measures reflect downstream metabolic consequences. Thus, deviation from the EPL should be interpreted as a complementary risk stratification marker that identifies structurally driven risk before overt biochemical failure becomes evident.

The observed associations between deviation metrics and measured visceral adiposity provide biological validation. Although bioelectrical impedance is not a gold standard, concordance with imaging-derived visceral fat supports the interpretation that EPL deviation reflects intra-abdominal fat predominance. Several anthropometric indices address BMI’s limitations, including the waist-to-height ratio, the body shape index, the body roundness index, and WWI. (10-13) While these indices incorporate central adiposity to varying degrees, they rely on fixed thresholds or lack a physiological reference state. In contrast, the iBMI–EPL framework enables deviation-based interpretation across a continuous spectrum, independent of absolute cut-offs. This framework aligns with contemporary calls to move beyond BMI toward metabolically informed definitions of obesity. (14) By integrating waist circumference into a height-neutral formulation and interpreting deviations from a physiological reference rather than fixed thresholds, the iBMI–EPL approach addresses a diagnostic blind spot that precedes overt dysglycemia. Longitudinal evidence indicates that substantial β-cell functional decline occurs before traditional biochemical thresholds are crossed, underscoring the need for upstream risk markers. (15)

The accompanying nomogram translates this concept into a clinically intuitive tool that enables visualization of deviation-based risk using routine anthropometric measurements. GRD zoning facilitates pragmatic stratification while preserving the continuous signal. These zones are operational aids and should be interpreted alongside clinical and biochemical factors. To support clinical implementation, a publicly accessible web-based calculator was developed to compute iBMI, deviation metrics, and graphical risk distance from routine anthropometric inputs, visualize results on the iBMI–BMI nomogram, and, optionally, store results for longitudinal documentation. (16) For contextual interpretation, the calculator allows entry of population-specific reference values; in Indian populations, a nominal normal iBMI of approximately 35 has been derived from anthropometric data. (17)

Several limitations warrant consideration. The retrospective, clinic-based design may limit generalizability, and sex-specific effects were not examined. Visceral fat imaging was available only in a subset. Prospective studies are needed to determine whether deviation-based metrics predict incident diabetes or cardiometabolic outcomes and to refine population-specific calibration. As with all observational analyses, residual confounding cannot be ruled out.

## Conclusion

This study proposes an integrated anthropometric framework that extends beyond body mass index by incorporating waist circumference within a height-neutral formulation and anchoring interpretation to an empirically derived adipose tissue equipoise line. Deviation from this reference state, quantified as the graphical risk distance or its algebraic equivalent, provides a continuous, physiologically grounded measure of central dysmetabolic adiposity rather than a categorical surrogate for body size alone. By distinguishing anatomically driven adipose imbalance from downstream glycemic or insulin-related abnormalities, this approach addresses a diagnostic blind spot that precedes overt biochemical deterioration. The iBMI–EPL framework enables visualization and quantification of compensated yet dysmetabolic states that are frequently overlooked by conventional indices, thereby facilitating earlier recognition of risk, counseling, and preventive intervention before established cardiometabolic disease develops. The accompanying nomogram and calculator translate this concept into a clinical tool using routine measurements, supporting individualized risk stratification while remaining adaptable to population-specific norms.

## Data Availability

All data produced in the present study are available upon reasonable request to the authors.

## Ethical approval

The study was conducted in accordance with institutional and international ethical standards. The ethics committee of Poona Hospital & Research Center, Pune, India, issued a waiver in accordance with the ICMR National Ethical Guidelines (2017).

## Acknowledgements

The authors have no acknowledgments to declare.

## Declaration of generative AI and AI-assisted technologies in the writing process

During manuscript preparation, the authors used ChatGPT (OpenAI) solely to assist with language editing and structural refinement; responsibility for the content rests entirely with the authors.

## Declaration of competing interest

The authors declare that they have no known competing financial interests or personal relationships that could have influenced the work reported in this paper.

**CRediT authorship contribution statement**

S.N.S.: Conceptualization, Data curation, Formal analysis, Writing–original draft. R.S.S.: Methodology, Visualization, Writing–review & editing. S.Y.B.: Clinical data collection, Supervision, Resources, Writing–review & editing.

S.N.S. is the guarantor of this work and, as such, had full access to all the data in the study and takes responsibility for the integrity of the data and the accuracy of the data analysis.

## Reporting Guideline Statement

### Reporting Guideline Statement

This study is a retrospective, cross-sectional observational analysis and is reported in accordance with the STROBE (Strengthening the Reporting of Observational Studies in Epidemiology) guidelines. The completed STROBE checklist is provided as supplementary material.

### Data availability statement

The datasets generated during and/or analyzed during the current study are available from the corresponding author on reasonable request.

